# Evaluating Effects of Resting-State Electroencephalography Data Pre-Processing on a Machine Learning Task for Parkinson’s Disease

**DOI:** 10.1101/2023.03.06.23286826

**Authors:** Robin Vlieger, Elena Daskalaki, Deborah Apthorp, Christian J. Lueck, Hanna Suominen

**Author notes:** Corresponding Author: Robin Vlieger, Hanna Neumann Building, Australian National University, 145 Science Road, Acton ACT 2601.

## Abstract

Resting-state electroencephalography (RSEEG) is a method under consideration as a potential biomarker that could support early and accurate diagnosis of Parkinson’s disease (PD). RSEEG data is often contaminated by signals arising from other electrophysiological sources and the environment, necessitating pre-processing of the data prior to applying machine learning methods for classification. Importantly, using differing degrees of pre-processing will lead to different classification results. This study aimed to examine this by evaluating the difference in experimental results when using re-referenced data, data that had undergone filtering and artefact rejection, and data without muscle artefact. The results demonstrated that, using a Random Forest Classifier for feature selection and a Support Vector Machine for disease classification, different levels of pre-processing led to markedly different classification results. In particular, the presence of muscle artefact was associated with inflated classification accuracy, emphasising the importance of its removal as part of pre-processing.

## 1. Introduction

Resting-state electroencephalography (RSEEG) is a method under consideration as a potential biomarker that could support early and accurate diagnosis of Parkinson’s disease (PD),[1, 2] as it can differentiate people with PD (PwPD) from controls.[3] In an effort to create diagnostic models, researchers have applied machine learning (ML) methods:[4] in most cases, RSEEG data are represented as features (e.g. power or entropy), followed by classifying each person’s feature vector using either conventional ML algorithms, such as Support Vector Machines (SVMs) and Random Forest Classifiers (RFCs), or deep learning approaches. Classification accuracy has ranged from 62.0% to 99.9%, with most experiments focusing on differences between controls and PwPD, though some have looked at the severity of cognitive impairment.[4]

Unfortunately, there have been large differences in the pre-processing approaches in different studies: some simply process raw data, some apply filters, and some use full pre-processing pipelines that include filtering, removal and interpolation of electrodes that are contaminated with noise, and artefact rejection.[4] This makes comparison and, indeed, interpretation of the results difficult. For example, inclusion of muscle artefact in the signal usually leads to higher classification accuracy,[5] but this implies that the classification was, at least partially, based on non-neural activity.

The goal of this study was therefore to evaluate the difference in classification performance resulting from different pre-processing pipelines. Features were created by calculating absolute and relative power at three stages: after re-referencing, after running a full pre-processing pipeline that included filtering, electrode removal and interpolation, and artefact rejection, and after running the same pipeline but without removing muscle artefact. We used three independent data sets for our experiments.

## 2. Methods

Three data sets were used for this study (Table 1). One collected by direct measurement at The Canberra Hospital (TCH) in Australia, and two were publicly available data sets provided by the University of New Mexico (UNM), USA, and the University of Turku (UTU), Finland.[1] Data were pre-processed using MATLAB R2018b and the MATLAB toolbox EEGLAB 2020.0,[6] following the recommendations of the Swartz Centre of Computational Neuroscience and Makoto Miyakoshi.[7] Data were re-referenced to the average of the electrodes and down-sampled to 128 Hz. A 1 Hz high-pass filter and 50 Hz low-pass filter were applied, and noisy channels were removed and interpolated using the EEGLAB extension ‘clean_rawdata’. Non-stationary artefacts were removed with the Artifact Subspace Reconstruction (ASR) function of ‘clean_rawdata’. Independent Component Analysis (ICA) and IClabel[8] were used to identify and remove independent components (ICs) that were more than 70% likely to not be of neural origin. Six regions of interest (ROIs) over the scalp were created by averaging electrodes based on location, and the signal was divided into 2-second epochs that were averaged for each ROI. Features, based on those used in the literature[4], were extracted after re-referencing, after filtering, after running the full pre-processing pipeline, and after running the full pipeline without removing muscle artefact. Absolute power in each of six frequency bands - delta, theta, alpha, alpha1 (8-10 Hz), alpha2 (10-13 Hz), and beta -was extracted using a Fourier transformation at each ROI, and these values were then used to calculate relative power, i.e. the alpha1-to-theta ratio and the ratio between alpha-plus-theta and alpha-plus-beta. In total, there were 84 features.

**Table 1.** Details of the three data sets used in this study.

| Data set | Recording Duration | N PwPD (female, male) | N Controls (female, male) | PwPD mean age [years] | Controls mean age [years] |
| --- | --- | --- | --- | --- | --- |
| TCH | 3 minutes | 21 (13, 8) | 18 (9, 9) | 68.2 ± 9.2 | 68.4 ± 8.7 |
| UNM | 1 minutes | 28 (11, 17) | 28 (11, 17) | 69.8 ± 8.6 | 69.2 ± 9.2 |
| UTU | 2 minutes | 20 (11, 9) | 20 (12, 8) | 70.0 ± 7.2 | 68.0 ± 6.0 |

An SVM classifier with a radial basis function kernel was used for the classification. Data were normalised using PowerTransformer.[9] Feature selection was performed using an RFC, only retaining the top 30 features based on Gini-coefficient, during 10-fold cross-validation repeated 10 times. Hyperparameters of the SVM were optimised, while the optimal number of features was obtained by starting out with the 2 best features, finding the best hyperparameters, then adding the next best feature and optimising performance again, until all features were included in the model. Performance was evaluated using accuracy, sensitivity (or recall), specificity, precision (also known as the positive predictive value), and F1-score, and significance tests were performed using Welch’s t-tests and a Bonferroni-corrected alpha level of .0167. All experiments were implemented in Scikit-learn in Python 3.8.[9]

For cross-validation, we used a train/validation split without a test set. This was necessitated by the small sizes of the data sets and warranted because the aim of the study was to evaluate the effects pre-processing on an ML classification task rather than assessing the generalisation capabilities of the ML classifier on unseen data. The “best-performing” model should be based on neural, rather than non-neural signals, and the test set would suffer even more severely from the same problem, as the data would be pre-processed in the same manner.

Collection of the TCH data set and use of all three data sets for this study were approved by the ACT Health Human Research Ethics Committee (ETH.4.16.060) and the Australian National University (ANU) Human Research Ethics Committee (protocol No. 2020/612). Written consent was obtained from all participants in TCH data set.

## 3. Results

Comparison of accuracy results of full pre-processing to full pre-processing retaining muscle artefact using Welch’s t-tests showed a significant difference for the UNM and UTU data sets (p < 0.0001 for both), accuracy being better if muscle artefact was included (Table 2). Features selected by the RFC differed between the fully pre-processed data and data with muscle artefact, as well as between data sets, and were very diffuse, so features were grouped by ROI and frequency band (Tables 3 and 4). For example, relative alpha and absolute alpha were grouped together. For the TCH data, 4 features were selected for the fully pre-processed data and 7 for data with muscle artefact, while for the UNM and UTU data sets these numbers were 2 and 30, and 29 and 4, respectively.

**Table 2.** Evaluation metrics from the described experiments in percentages on the validation set. ‘Full’ refers to the full pre-processing pipeline described in the methods and ‘Full, retaining artefact’ refers to full pre-processing with muscle artefact left in. The best scores for each data set are in bold.

| Data set | Pre-processing stage | Accuracy ± SD | Sensitivity | Specificity | Precision | F1-score |
| --- | --- | --- | --- | --- | --- | --- |
| TCH | Re-referenced | 65.13±24.43 | 70.48 | 58.89 | 66.67 | 68.52 |
| | Filtered | 58.97±24.67 | 60.48 | 57.22 | 62.25 | 61.35 |
| | Full, retaining artefact | 69.74±19.07 | **71.43** | 67.78 | 72.12 | 71.77 |
|  | Full | **73.85±21.20** | 65.71 | **83.33** | **82.14** | **73.02** |
| UNM | Re-referenced | 66.96±17.36 | 61.79 | 72.14 | 68.92 | 65.16 |
|  | Filtered | **85.00±13.89** | **83.93** | **86.07** | **85.77** | **84.84** |
| | Full, retaining artefact | 79.11±15.10 | 75.00 | 83.21 | 81.71 | 78.21 |
| | Full | 66.07±20.36 | 57.86 | 74.29 | 69.23 | 63.04 |
| UTU | Re-referenced | 52.31±25.48 | 51.05 | 53.50 | 51.05 | 51.05 |
| | Filtered | 57.44±25.46 | 58.42 | 56.50 | 56.06 | 57.22 |
|  | Full, retaining artefact | **71.03±21.52** | **63.68** | **78.00** | **73.33** | **68.17** |
| | Full | 51.79±23.73 | 47.37 | 56.00 | 50.56 | 48.91 |

**Tables 3 and 4.**
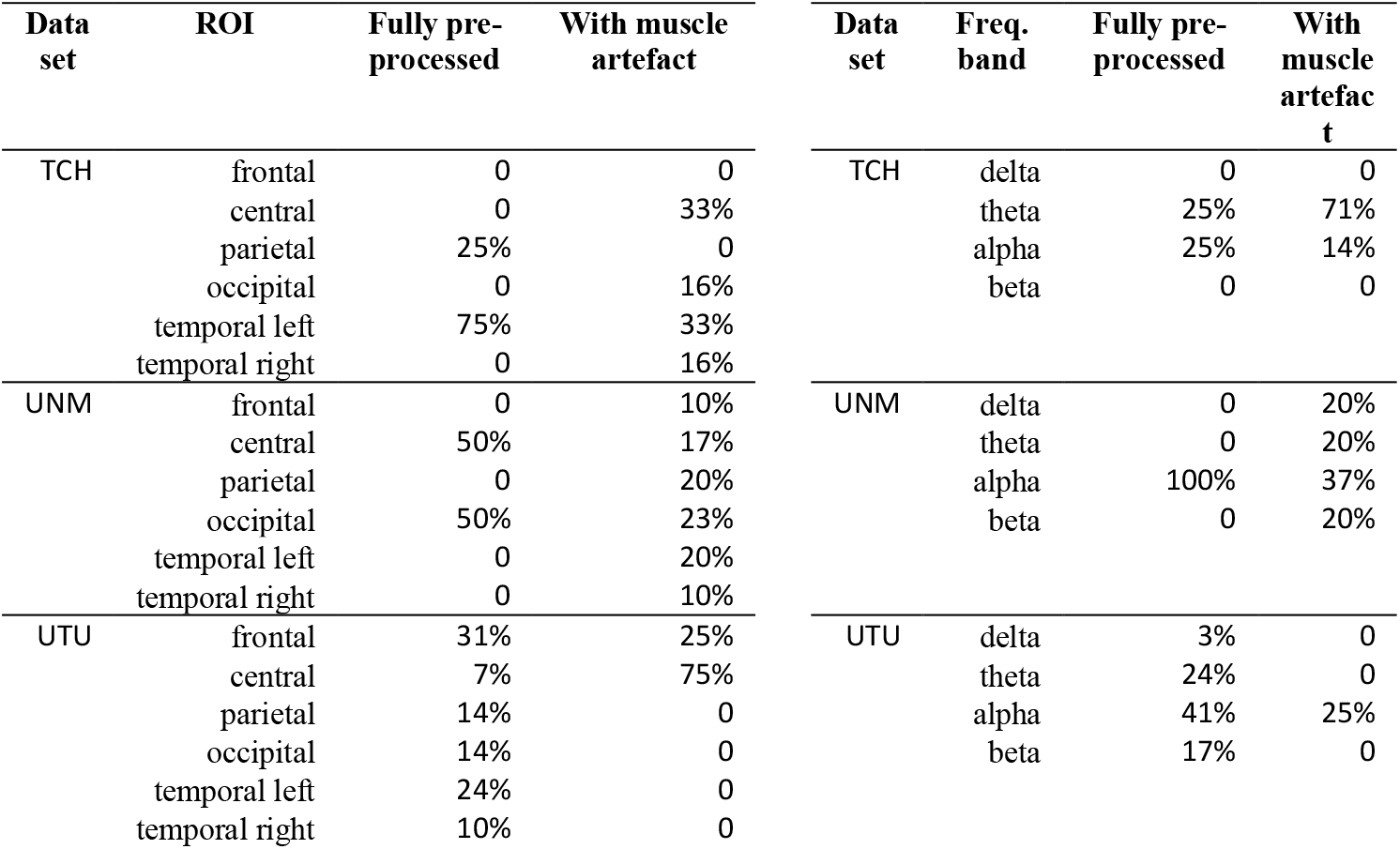
Distribution of features across ROIs and per frequency band, per data set, and per pre-processing stage, expressed as a percentage of total number of features selected by the RFC. Note that frequency band percentages do not always add up to 100% as they do not include the ratio features.

| Data set | ROI | Fully pre-processed | With muscle artefact | Data set | Freq. band | Fully pre-processed | With muscle artefact |
| --- | --- | --- | --- | --- | --- | --- | --- |
| TCH | frontal | 0 | 0 | TCH | delta | 0 | 0 |
|  | central | 0 | 33% |  | theta | 25% | 71% |
|  | parietal | 25% | 0 |  | alpha | 25% | 14% |
|  | occipital | 0 | 16% |  | beta | 0 | 0 |
|  | temporal left | 75% | 33% |  |  |  |  |
|  | temporal right | 0 | 16% | UNM | delta | 0 | 20% |
| UNM | frontal | 0 | 10% |  | theta | 0 | 20% |
|  | central | 50% | 17% |  | alpha | 100% | 37% |
|  | parietal | 0 | 20% |  | beta | 0 | 20% |
|  | occipital | 50% | 23% |  |  |  |  |
|  | temporal left | 0 | 20% |  |  |  |  |
|  | temporal right | 0 | 10% | UTU | delta | 3% | 0 |
| UTU | frontal | 31% | 25% |  | theta | 24% | 0 |
|  | central | 7% | 75% |  | alpha | 41% | 25% |
|  | parietal | 14% | 0 |  | beta | 17% | 0 |
|  | occipital | 14% | 0 |  |  |  |  |
|  | temporal left | 24% | 0 |  |  |  |  |
|  | temporal right | 10% | 0 |  |  |  |  |

## 4. Discussion

We evaluated the differences in classification performance of PwPD and controls when using different levels of pre-processing of RSEEG data as used by studies in the literature[4] To enhance the validity of our observations, we used three different data sets and investigated the differences in features selected to attain these performance metrics. The highest metrics were achieved for the TCH data set using fully pre-processed data, for the UNM data set using filtered data, and for the UTU data set using data with muscle artefact. Of note, for the UNM and UTU data sets, classification metrics were improved when using data retaining muscle artefact than when using fully pre-processed data.

Interestingly, contrary to the other data sets, fully pre-processing the TCH data set increased performance. There are several possible explanations for this. For example, the EEG data were noisy, and the data sets were small (39, 43, and 56 total participants in the TCH, UNM, and UTU data sets, respectively). It is also possible that the controls in the TCH data set had more muscle artefact in their recordings compared to the other data sets, which obscured the signal and was filtered out, leading to improvements.

The best-performing ROIs were not consistent across data sets. However, feature analysis was clearer: using fully pre-processed data, absolute power in the theta band (4-8 Hz) contributed most for the TCH and UTU data sets, while absolute power in the alpha1 band contributed most for the UNM data set. Of note, the cut-off points between frequency bands are still being debated[10,11], so two data sets being dominated by theta features and one data set by alpha1 features was not too surprising. Grouping frequency bands together would not be unreasonable and this suggests that a more detailed analysis of frequency in the future research is worthwhile.

## 5. Conclusion

Our results indicate that removal of artefacts is essential if the intention is to classify subjects based on neural activity. When this is done, theta and alpha features contribute most to classification accuracy. Further research is needed to determine which specific features are necessary for accurate classification.

## Data Availability

The data collected at The Canberra Hospital in the present study are available upon reasonable request to the authors.
The data collected at the University of Turku are available online at https://osf.io/pehj9/
The data collected at the University of New Mexico are available at http://predict.cs.unm.edu/downloads.php

http://predict.cs.unm.edu/downloads.php

## 6. Acknowledgment

This research was funded by and has been delivered in partnership with Our Health in Our Hands (OHIOH), a strategic initiative of the Australian National University, which aims to transform health care by developing new personalized health technologies and solutions in collaboration with patients, clinicians, and health-care providers. We gratefully acknowledge the funding from the ANU School of Computing for the first author’s PhD studies.

